# Adeno-associated virus type 2 in children from the United States with acute severe hepatitis

**DOI:** 10.1101/2022.09.19.22279829

**Authors:** Venice Servellita, Alicia Sotomayor Gonzalez, Daryl M. Lamson, Abiodun Foresythe, Hee Jae Huh, Adam L. Bazinet, Nicholas H. Bergman, Robert L. Bull, Karla Y. Garcia, Jennifer S. Goodrich, Sean P. Lovett, Kisha Parker, Diana Radune, April Hatada, Chao-Yang Pan, Kyle Rizzo, J Bradford Bertumen, Christina Morales, Paul E. Oluniyi, Jenny Nguyen, Jessica Tan, Doug Stryke, Rayah Jaber, Matthew T. Leslie, Zin Lyons, Hayden Hedman, Umesh Parashar, Maureen Sullivan, Kelly Wroblewski, M. Steven Oberste, Jacqueline E. Tate, Julia M. Baker, David Sugerman, Caelin Potts, Xiaoyan Lu, Preeti Chhabra, CDC Pediatric Hepatitis of Unknown Etiology Working Group, L. Amanda Ingram, Henry Shiau, William Britt, Luz Helena Gutierrez Sanchez, Caroline Ciric, Christina A. Rostad, Jan Vinjé, Hannah L. Kirking, Debra A. Wadford, R. Taylor Raborn, Kirsten St. George, Charles Y. Chiu

## Abstract

As of August 2022, cases of acute severe hepatitis of unknown etiology in children have been reported from 35 countries, including the United States. Here we used PCR testing, viral enrichment based sequencing, and agnostic metagenomic sequencing to analyze 27 samples, including nasopharyngeal swab, stool, plasma, and/or whole blood, from 16 such cases from 6 states (Alabama, California, Florida, Illinois, North Carolina, and South Dakota) presenting from October 1, 2021 to May 22, 2022, in parallel with whole blood samples from 45 controls. Among the 13 cases for whom whole blood was available, adeno-associated virus 2 (AAV2) sequences were detected in 92% (12 of 13) (p<0.001), while adenovirus sequences were detected in 100%, of which 11 (84.6%) were genotyped as human adenovirus type 41 (HAdV-41), one as HAdV-40, and one as HAdV-2. Co-infections of herpesviruses, Epstein-Barr virus (EBV) or human herpesvirus 6 (HHV-6), and/or enterovirus A71 (EV-A71) were also detected in all 13 cases. In contrast, AAV2 and HAdV-41 were not detected in any control, and EBV, HHV-6, or EV-A71 were each only detected in one or two of 45 controls (p<0.001). Analysis of assembled AAV2 viral genome sequences identified 35 coding mutations relative to the AAV2 reference genome, predominantly located in the VP1 capsid and assembly-activating protein (AAP) proteins, and AAV2 genomes from cases clustered together by phylogenetic analysis. Our findings of a distinct AAV2 strain in nearly all cases of acute severe hepatitis of unknown etiology in conjunction with one or more infecting helper viruses suggest that disease pathogenesis and/or severity may be related to co-infection with AAV2.

## INTRODUCTION

Viral hepatitis is an inflammation of the liver most commonly caused by one of the major hepatitis viruses (A-E). Since October 2021, outbreaks of acute non A-E severe hepatitis of unknown etiology in children have been reported in 35 countries, including the United States (US)^1,2^. The majority of pediatric cases globally have been <6 years of age (76%), with an approximately equal gender distribution (48% male to 52% female)^2^. As of August 17, 2022, 358 persons under investigation (PUIs) have been reported in the United States, of whom 22 (6%) required a liver transplant and 13 (4%) died^3^.

Human adenoviruses (HAdVs) are non-enveloped, double-stranded DNA viruses that cause a variety of infections in both adults and children, including respiratory tract infection, conjunctivitis, and gastroenteritis (mainly from adenovirus species F including HAdV types 40 and 41^4,5^. Although previously reported as a severe and potentially fatal disease in immunocompromised patients^4,6^, hepatitis from adenovirus infection had been thought to be rare in immunocompetent children without underlying comorbidities^7^. In previous reports, adenoviruses, particularly adenovirus type 41 (HAdV-41), have been found in the blood from a majority of outbreak-associated cases of acute severe hepatitis from Scotland, the United Kingdom (UK), and the US^6,8-11^, although it is unclear whether this virus is causative. AAVs have also been reported in children with severe acute hepatitis in a study of 9 patients from the United Kingdom (UK)^18^, often in association with adenovirus or human herpesvirus 6 (HHV-6 infection)^12^. AAVs are small, single-stranded DNA parvoviruses that are thought to be non-pathogenic in humans and for this reason have been widely used as vectors for gene therapy^13^. Importantly, AAVs require a helper virus, such as a herpesvirus or adenovirus, for productive infection, although the contribution of AAVs to hepatitis remains unclear^14,15^.

In this study, we employ a variety of molecular and sequencing-based methods to investigate acute viral infections that may be causing severe hepatitis in children. These include metagenomic sequencing for agnostic detection of all viruses^16^, tiling multiplex PCR amplicon sequencing for AAV2 and HAdV-41^17^, metagenomic sequencing with probe capture viral enrichment for 3,153 viruses^18^, and virus-specific polymerase chain reaction (PCR). Our findings in affected US children from 6 geographically dispersed states demonstrate co-infection from a distinct strain of adeno-associated virus type 2 (AAV2) in nearly all cases and none of the controls.

## Results

### Patient and sample characteristics

In this study, 27 samples (21 whole blood, 2 plasma, 1 liver tissue, 1 nasopharyngeal swab, and 2 stool sample(s)) from 16 pediatric patients with acute severe hepatitis of unknown etiology were analyzed **(Figure 1A)**. All patients met the clinical case definition for the disease established by the CDC, including lack of a confirmed etiology for the hepatitis (including negative testing for hepatitis viruses A-E), liver enzyme levels (aspartate aminotransferase / AST or alanine aminotransferase / ALT) >500 UL, age <10 years, and onset on or after October 1, 2021^2,19^. Affected children were admitted to tertiary care hospitals in 6 states (Alabama, California, Florida, Illinois, North Carolina, and South Dakota) from October 1, 2021 to May 22, 2022 **(Extended Data Table 1)**. Because many patients (12 of 16, 75%) were selected from those PUIs who tested positive for adenovirus and had samples submitted for adenovirus typing, PUIs with adenovirus infection may have been over-represented compared to the overall PUI population, of which adenovirus is detected in 45-90%^6,8-11^. The median age of affected children was 3 years. Mean elevations in AST and ALT were 2,652 +/- 1,851 and 2,293 +/- 1,486, respectively; 2 children underwent liver transplantation and 0 died from complications of liver failure; the others are still hospitalized at time of data collection or recovered with supportive care.

**Figure 1.**
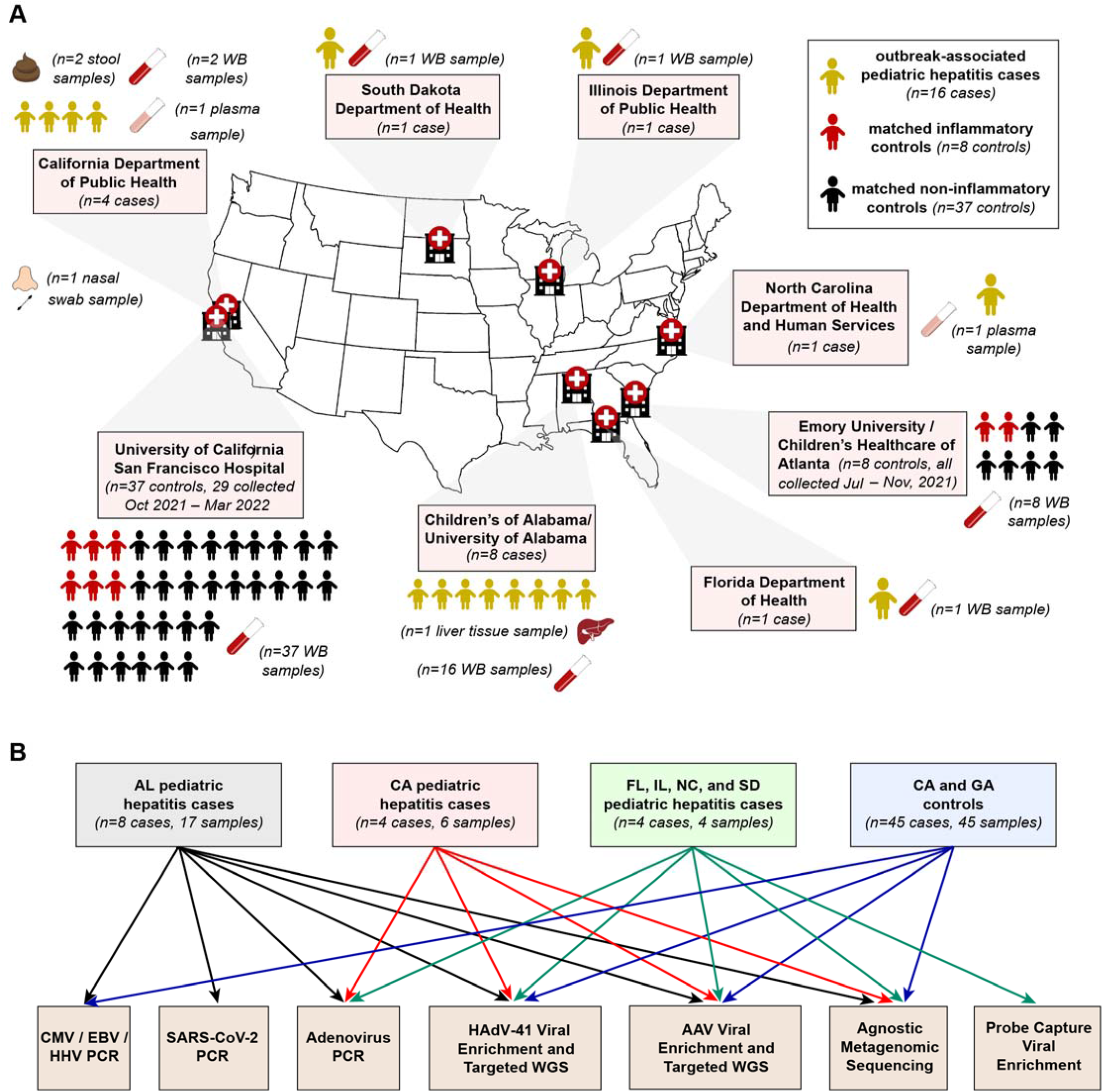
Epidemiology of acute severe hepatitis cases and controls and analyses performed on collected samples. **(A)** Geographic distribution of the 16 cases and 45 controls in the study. Controls were patients hospitalized with other inflammatory or non-inflammatory conditions or healthy blood donors. **(B)** Analyses performed on the different sample groups. Color-coded arrows indicate the specific assays that were performed for each cohort.

Whole blood samples from 45 pediatric controls, 8 from Georgia to be geographically similar (located in a neighboring state) to the Alabama and Florida cases, and 37 from California to be geographically similar (located in the same state) to the California cases, were analyzed in parallel. Geographically similar samples for the North Carolina, Illinois, and South Dakota cases were not available. Control samples were collected between August 4, 2020 and March 28, 2022. Of the 45 control samples, 23 (62%) were also collected over the same time frame as the cases (i.e., collected between October 1, 2021 and March 28, 2022). The median age of controls was 9 +/- 5 years. Among the 45 controls, 8 (18%) were hospitalized patients with a non-hepatitis inflammatory condition (e.g., sepsis, viral respiratory infection, osteomyelitis, etc.), 31 (69%) were hospitalized patient with a noninflammatory condition, while 6 (13%) were healthy blood donors.

### Detection of viruses in acute severe hepatitis cases

Metagenomic sequencing, tiling multiplex PCR amplicon sequencing, metagenomic sequencing with probe capture viral enrichment (VE), and virus-specific PCR were performed to identify viruses in clinical samples from acute severe hepatitis cases **(Figure 1B)**. We detected AAV2 from available whole blood samples in 12 (92%) of 13 cases and HAdV in all 13 cases **(Figure 2)**. Ten (77%) of the 13 cases were HAdV-41, one (7.7%) was HAdV-40, and one (7.7%) was HAdV-2. AAV2 and HAdV-41 enrichment were performed using either tiled multiplex PCR amplicon sequencing targeting the genomes of HAdV-41 and AAV (n=9 samples, with a mean 6.6±1.5 (SD) million raw reads generated per sample) or probe capture VE targeting 3,153 viral species (n=4, with a mean 128±34 (SD) million raw reads generated per sample). Mean viral read counts for AAV2 (6285 and 56815 reads from targeted and VE sequencing, respectively) were approximately 30-100 times greater than for HAdV-41 (58 and 1676 reads from targeted and VE sequencing, respectively) **(Extended Data Table 1)**. AAV2 was detected in both liver tissue and whole blood from one case and in 1 of 2 plasma samples, but neither AAV2 nor HAdV-41 were detected in nasopharyngeal swab and stool samples. No reads from AAVs other than AAV2 were detected, despite using methods that would enrich for other AAV subtypes.

**Figure 2.**
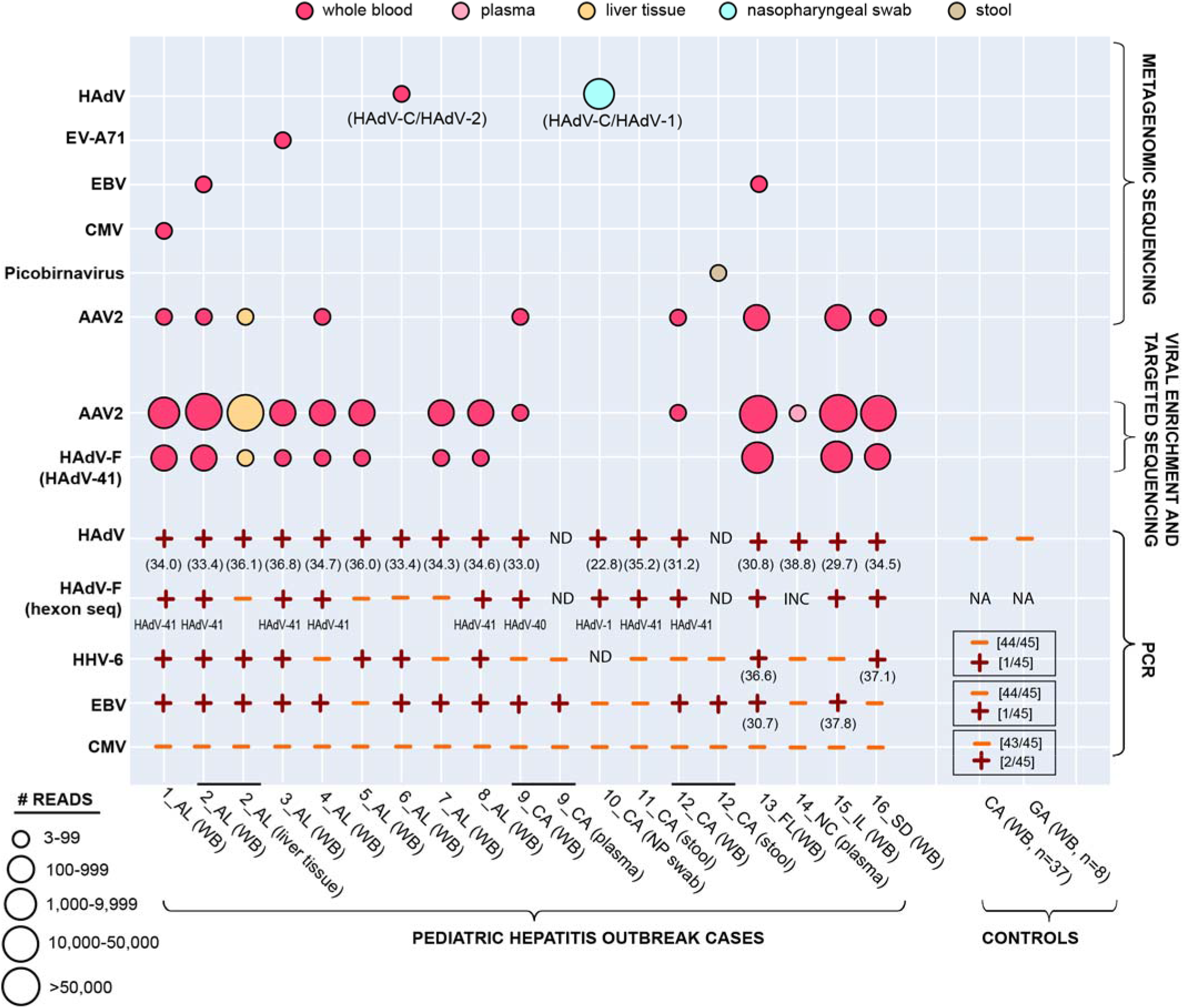
Overview of next-generation sequencing and molecular-based testing performed on cases and controls. Each circle represents a sequenced sample with at least 3 non-overlapping reads^20^ aligning to a viral sequence in the National Center for Biotechnology Information (NCBI) database detected by metagenomic and targeted or virally enriched sequencing. The circles are color-coded based on the sample type and size-scaled based on viral read counts. PCR positive and negative results are denoted by plus and minus symbols, respectively; with cycle threshold values listed under the plus symbol for HAdV-positive samples. Abbreviations: ND, not done; HAdV-C, human adenovirus type C; HAdV-F, human adenovirus type F; EV-A71, enterovirus A71; EBV, Epstein-Barr virus; CMV, cytomegalovirus; HAdV-41, human adenovirus type 41, HAdV-40, human adenovirus type 40, HHV-6, human herpesvirus 6; AAV, adeno-associated virus; AL, Alabama; CA, California; FL, Florida; NC, North Carolina; IL, Illinois; SD, South Dakota; GA, Georgia; PCR, polymerase chain reaction.

Metagenomic sequencing was less sensitive than viral enrichment and targeted sequencing for detection of AAV2 (8 of 13 cases, 61.5%) and HAdV-41 (0 of 13 cases, 0%) **(Figure 2 and Extended Data Table 1)**. However, reads from additional viruses were identified including EBV (n=2), CMV (n=1), HAdV-2 (adenovirus type C) (n=1), and enterovirus A71 (EV-A71) (n=1) from whole blood, HAdV-1 (adenovirus type C) (n=1) from nasopharyngeal swab, and picobirnavirus (n=1) from stool. EV-A71 was detected as a co-infection in an HAdV-41/AAV2 case, while HAdV-2 was detected in the single AAV2 negative case. Among the 45 whole blood controls, no viral pathogen was detected.

To identify additional viruses that may be associated with acute severe hepatitis, we performed virus-specific PCR testing for HAdV with genotyping by hexon gene sequencing, EBV, CMV, and HHV-6 for all cases and controls with sufficient volume for testing **(Figure 2 and Extended Data Table 1)**. AdV PCR confirmed the detection of HAdV-41 (n=11), HAdV-2 (n=1), and HAdV-1 (n=1), but also identified an additional case of HAdV-40 infection that was missed by targeted and metagenomic sequencing. EBV and HHV-6 were detected from whole blood in 11 (85%) of 13 and 8 (62%) of 13 cases, respectively, versus 1 (0.02%) of 45 controls for each virus. No tested cases were positive for CMV as compared to 2 (0.04%) of 45 controls.

Combining all 3 detection modalities yielded a positive detection for AAV2 in whole blood samples from 12 (92%) of 13 cases and adenovirus, either HAdV-41 (n=10), HAdV-40(n=1), or HAdV-2 (n=1) in all 13 cases. The latter result is not surprising given that most patients with whole blood samples, 10 (76.9%) of 13, were known to be adenovirus positive by prior clinical testing. Co-infections with EBV and HHV-6 were also found in a large proportion of cases tested for these specific viruses, 11 (85%) of 13 for EBV and 8 (62%) of 13 for HHV-6. HAdV-1 was detected in a single patient for whom only nasopharyngeal swab sample was available. Of the remaining 3 cases of the 16 total for whom only a stool or plasma sample was available, 2 were negative for both AAV2 and adenovirus while 1 plasma sample was positive for AAV2 only.

### Associations between detected viruses and acute severe hepatitis cases

We used Fisher’s Exact Test on cases (n=13) and controls (n=45) to investigate associations between detected viruses in whole blood and acute severe hepatitis. For these analyses, we included data from agnostic metagenomic sequencing, PCR, targeted sequencing for HAdV-41 and AAV2, and probe capture viral enrichment-based sequencing **(Figure 3)**. Among viruses detected by agnostic metagenomic sequencing (HAdV-41, AAV2, EV-A71, EBV, CMV, and HAdV-2), only AAV2 (*P*<.001) and EBV (*P*=.047) had a significant correlation with acute severe hepatitis **(Figure 3)**. Using real-time PCR assays with probes specific for the detection of HHV-6, EBV and CMV, significant correlations were observed between HHV-6 (*P*<0.001) and EBV (*P*<0.001) **(Figure 3B)**. Using a combination of metagenomic sequencing, probe capture viral enrichment, and targeted HAdV-41 and AAV2 sequencing, significant correlations were observed between AAV2 infection and acute severe hepatitis (p<0.001) **(Figure 3C)**. A significant correlation was also observed between adenovirus infection and acute hepatitis (p<0.001), even after removal of the 10 (76.9%) of 13 cases that were known to be positive for adenovirus *a priori* **(Figure 3D)**.

**Figure 3.**
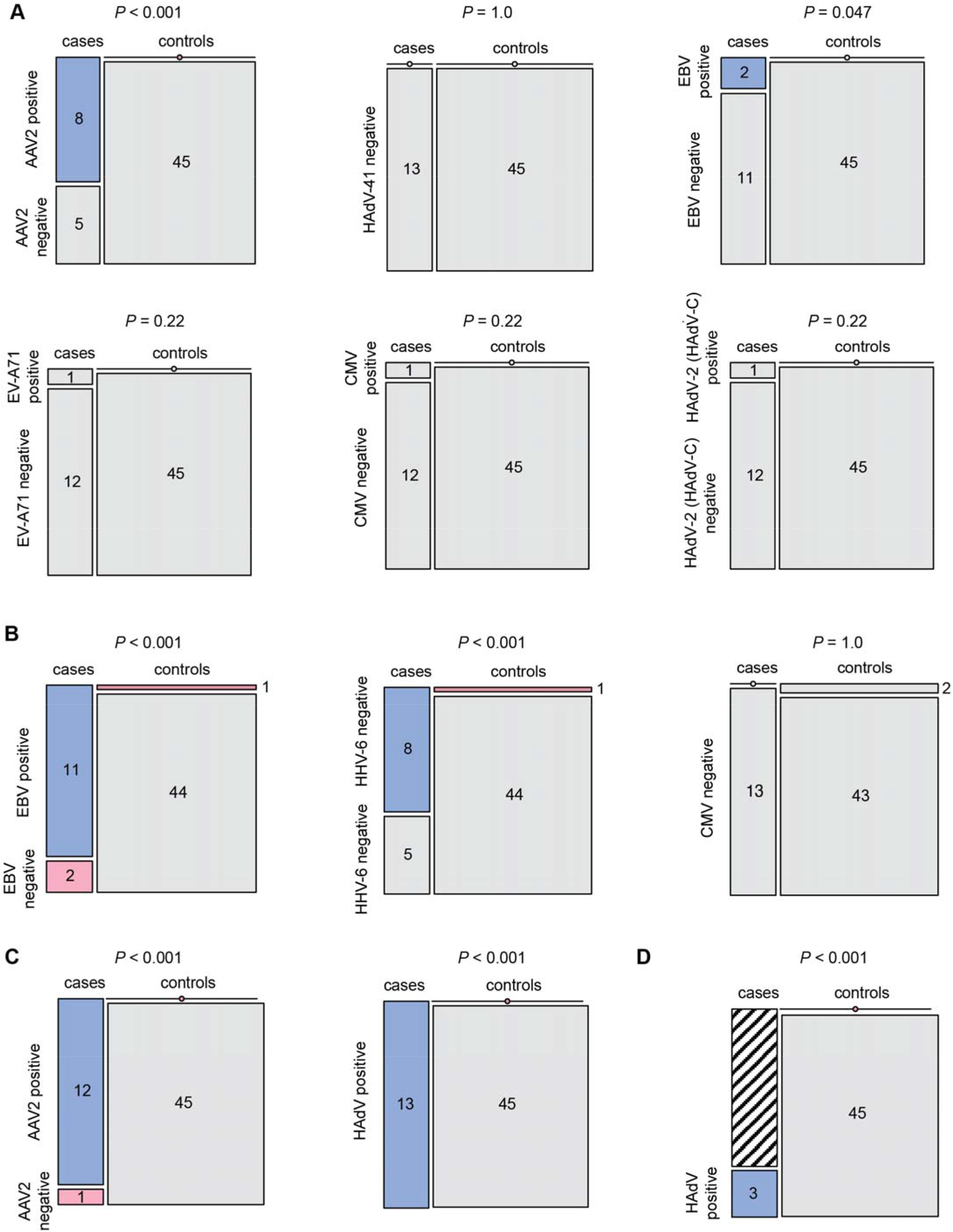
Associations between detected viruses in whole blood and cases of acute severe hepatitis of unknown etiology in children. **(A)** Virus detection by metagenomic next-generation sequencing. **(B)** Virus detection by specific PCR, **(C)** AAV2 detection by metagenomic, probe capture enrichment, and/or targeted multiplex PCR amplicon sequencing. **(D)** HAdV detection by metagenomic, probe capture enrichment, and/or targeted multiplex PCR amplicon sequencing, before (left) and after (right) removal of the 10 (76.9%) of 13 cases known to be positive for adenovirus by *a priori* clinical testing. Fisher’s Exact test (two-tailed) was used to calculate *P* values.

### Phylogenetic and mutation analyses

Fourteen complete or partial AAV2 genomes were recovered from 12 whole blood samples, one liver tissue, and one plasma from 13 cases **(Figure 2)**. We performed multiple sequence alignment and phylogenetic analysis of 13 recovered AAV2 genomes from 12 cases with >25% breadth of coverage **(Figures 4 and 5)**. One AAV2 genome, corresponding to sample 14_NC, had <17% coverage so was excluded from further analysis. The multiple sequence alignment was performed in parallel with all complete 119 AAV2 reference genomes deposited in GenBank as of August 18, 2022, consisting of sequences from humans (n=108), non-human primates (n=30), geese and ducks (n=3), rodents (n=3), and bats (n=2). A nucleotide phylogenetic tree constructed from the genomes and rooted using a bat AAV genome (NC_014468.1) demonstrated clustering of the 13 recovered genomes from this study within a subgroup of a single large human AAV2 clade **(Figure 4A)**. Other previously sequenced AAV2 genomes from France (MK139269.1 and MK139259.1) and the United States (KY271943.1, AY695372.1, AY695374.1 and AY695375.1) from patients without hepatitis were also found within this subgroup. Amino acid phylogenetic trees of the VP1 and AAP proteins showed slightly different topologies, but the 13 recovered genomes continued to cluster together in a distinct AAV2 subgroup within one of the subclusters.

**Figure 4.**
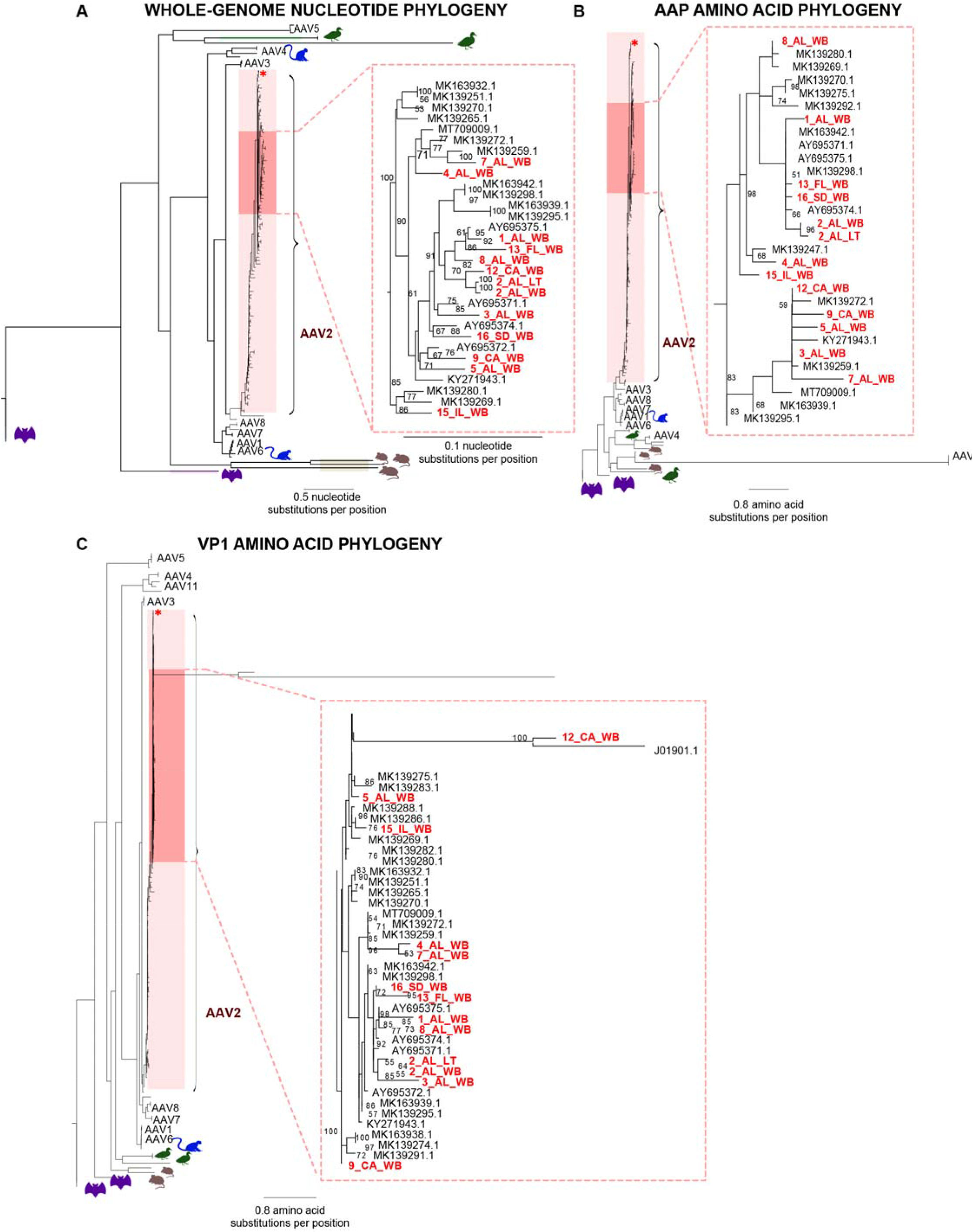
Phylogenetic tree of AAV2 virus from the acute hepatitis outbreak of 2022. **(A)** Phylogenetic tree of 119 AAV2 whole genomes available from GenBank as of August 18, 2022 and 12 recovered genomes from this study with >25% coverage denoted in red. **(B)** Phylogenetic tree from amino acids corresponding to the Assembly Activating Protein (AAP) protein. **(C)** Phylogenetic amino acid tree corresponding to the VP1 structural protein. The phylogenetic tree was constructed by multiple sequence alignment of the AAV genomes or amino acid sequences using the MAFFT algorithm^22^, followed by maximum likelihood based tree construction using IQ-TREE^23^. The location of the AAV2 reference genome (NC_001401.2) is marked with a red asterisk.

**Figure 5.**
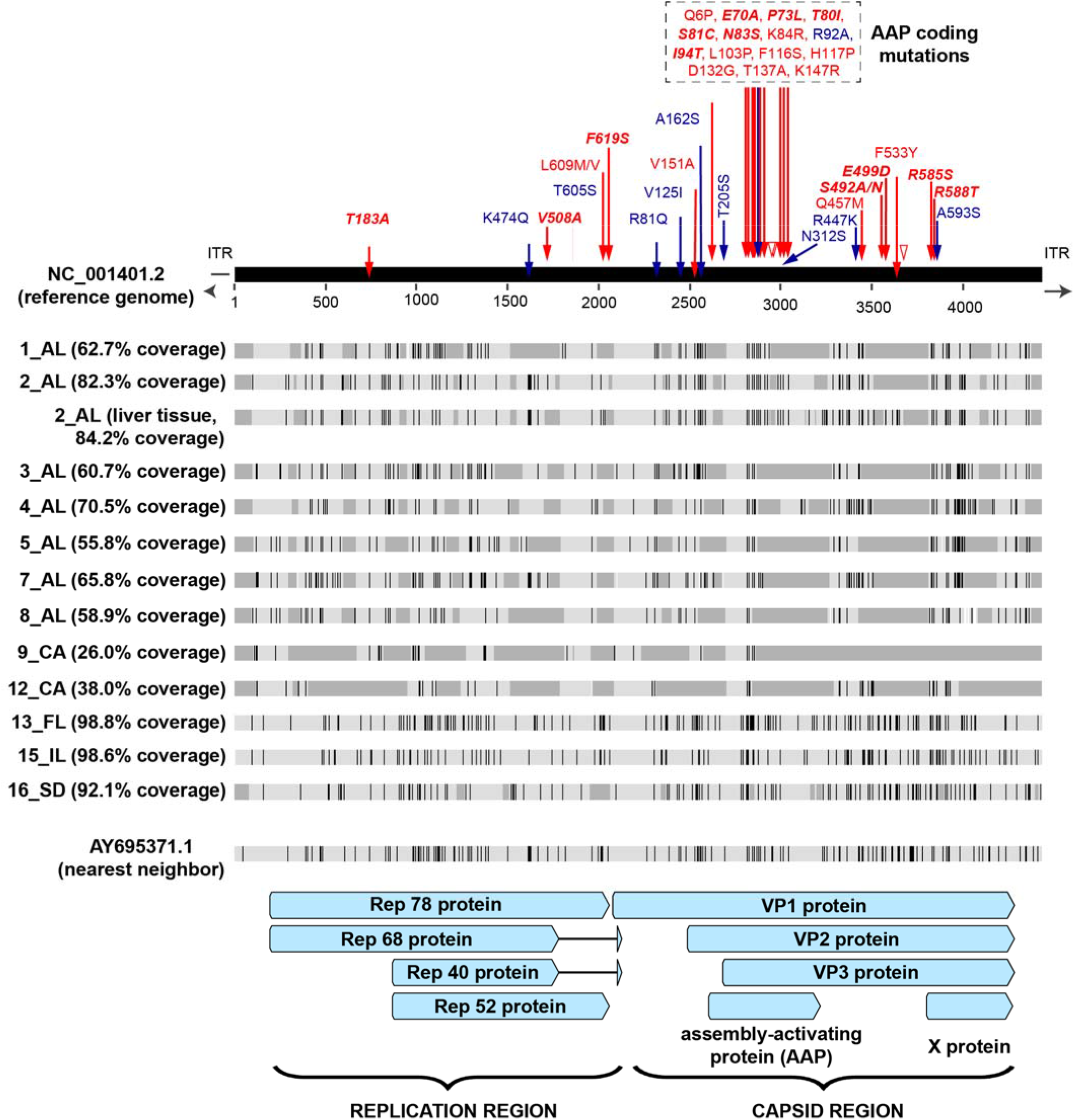
Adeno-associated virus 2 variant analysis. Multiple sequence alignment of adeno-associated virus genomes recovered from the acute hepatitis cases. Nucleotide mismatches are represented by black-colored vertical lines, while areas of missing coverage are represented by gray rectangles. Amino acid variants with respect to the reference genome (NC_001401.2) are denoted in blue and red arrows; red arrows indicate shared mutations that were reported in another study from the UK and Scotland^12^. Mutational sites that were identified in 100% of cases with sufficient coverage in both studies are highlighted in boldfaced red text.

To explore the underlying basis behind the phylogenetic clustering, we performed a multiple sequence alignment of the 13 recovered AAV2 genomes and searched for shared coding mutations relative to the AAV2 reference genome (NC_001401.2) that were found in ≥50% of recovered genomes with coverage at the nucleotide site. This analysis yielded 35 mutations that were unevenly distributed across the viral genome with 15 of 35 (42.9%) in the assembly activating protein (AAP), 14 of 35 (40%) in the VP1 protein, and 6 of 35 (17.1%) in the Rep78 protein **(Figure 5 and Extended Data Table 2)**. Clusters of mutations were found to be located within hypervariable regions of the capsid VP1 and AAP proteins^21^. Of note, 25 (71.4%) of 35 mutations were shared with those identified in an independent study of severe pediatric hepatitis from the UK^12^, with 12 (34.3%) of 35 mutations found in 100% of recovered AAV2 genomes (with coverage at the variable site) from both studies.

## Discussion

We report here virus findings from PCR, metagenomic, and targeted sequencing of samples from 16 pediatric cases of acute severe hepatitis of unknown etiology that were first identified in the United States in October 2021. After combining these three diagnostic modalities, AAV2 was detected in 92% (12 of 13) of cases for which whole blood was available (*P*<.001). HAdV sequences were detected in all 13 cases, of which 11 (84.6%) were genotyped as HAdV-41. Conversely, no control specimens had AAV2 and HAdV-41 detected. Although the 100% detection rate of HAdV may not be surprising as 10 (76.9%) of the 13 cases were known to be adenovirus positive by clinical testing *a priori*, the detection of HAdV in the 3 remaining cases and in 0 of 45 controls is still significant (p<0.001); further, adenovirus is detected in 45-90% of the overall PUI population^6,8-11^. Other co-infecting viruses, including EBV and HHV-6, were detected by PCR in many cases but rarely in controls (*P*<.001); EV-A71 was also detected by metagenomic sequencing in one case but not in any control. These findings indicate an association between co-infection by AAV2 and one or more hepatotropic viral pathogens and the clinical manifestations of severe acute hepatitis, although a direct causal link has yet to be confirmed.

Initial epidemiological investigation and molecular testing have found adenovirus in 45-90% of severe acute hepatitis cases^6,8-11^. In the UK, 116 of 179 (64.8%) reported cases as of May 2022 have tested positive for adenovirus^1^. This is in comparison to 8 of 9 (88.9%) positive cases in the United States corresponding to a cluster of cases who presented at a children’s hospital in Alabama between October 2021 and March 2022^10^. Slightly less than half (44.6%) of PUIs in an expanded national investigation in the United States had adenovirus detected^8^. In the current study, 9 (75%) of 13 cases were already known to be adenovirus positive *a priori* by clinical testing, with the remaining 4 cases adenovirus positive by viral PCR and/or sequencing. Consistent with prior reports^10,12^, genotyping by sequencing confirmed that most of the detected serotypes were HAdV-41 (10 of 13, 77%).

Our results suggest that co-infection with AAV2 may cause ta more severe liver disease than infection by an adenovirus and/or herpesvirus alone. This condition may be analogous to the fulminant liver failure that can occur when hepatitis D virus (HDV, “delta”) infection is superimposed on a chronic hepatitis B virus (HBV) infection^24^. Here we found that in cases with AAV2/HAdV co-infection, viral loads for AAV2 were likely higher than for HAdV, with higher positivity rates and a 2-3 log_10_ increase in the number of reads by targeted sequencing (mean AAV2 reads = 7,252 versus HAdV-41 reads = 67). This observation may be partially explained by previously published data showing that AAVs can suppress the replication of other hepatotropic viruses, including adenoviruses and herpesviruses^15,25^. Similarly, patients co-infected with HDV usually exhibit low HBV viral loads due to suppression of HBV replication by HDV-induced interferons^26^.

Among the AAVs, AAV2 is the most well-characterized AAV and has been shown to replicate to high titers in the liver and spleen^27^. Seroprevalence data from infants, children, and adults demonstrate that 30-80% of the general population is seropositive and that natural infection with AAV can occur at all ages^28^. However, there is a peak of AAV2 infection between the ages of 1 to 5 years old^29^, consistent with the observed distribution of ages of affected pediatric cases in the current study.

A whole-genome nucleotide phylogeny and amino acid phylogenies of the VP1 and AAP proteins of all sequenced human AAVs and representative nonhuman AAVs reveal that the recovered genomes from all cases cluster together in a distinct subclade of AAV2. The clustering is driven by groups of mutations located within hypervariable regions of the capsid VP1 and AAP proteins^21^. Notably, two of these mutations in the VP1 protein, R585S and R588T, are arginine-to-serine and arginine-to-threonine mutations that likely impact receptor binding as these residues are necessary for the interaction of AAV2 with its heparan sulfate proteoglycan receptor^30^. These are shared not only among the US genomes in the current study but also overlap substantially with the mutations found in an independent study of severe pediatric cases from the UK^12^. Interestingly, several of these shared capsid mutations (V151A, Q457M, S492A, E499D, F533Y, R585S, and R588T) are also found in a sublineage of AAV2 (AAVv66) that exhibits increased replication, virion stability, central nervous system transduction, and evasion of neutralizing antibodies relative to wild-type AAV2. However, as other contemporary AAV2 genomes are not readily available, it is unclear whether the relatedness in AAV2 genomes across geographically dispersed regions and to AAVv66 merely represents detection of the predominant global circulating strain.

In the current study, hepatotropic viruses other than HAdV, including EBV, HHV-6, and EV-A71, were detected, albeit in a smaller proportion of cases. Infection by HHV-6 or EBV alone has been implicated in cases of liver failure requiring transplantation^31,32^. Notably, among the 13 cases, dual or triple infections with adenovirus and one or both herpesviruses were detected in whole blood from at least 11 cases (>85%). We postulate that the COVID-19 pandemic and more than 2 years of school and childcare closures, social distancing measures, and decreased overall social interactions may have generated a vulnerable population of young children who have failed to develop broad immunity to common viral pathogens due to lack of exposure. This may potentially explain the increased proportion of cases with multiple viral infections observed in the current study, including from AAV2, that we speculate may have increased the likelihood of more severe disease manifestations such as hepatitis.

In summary, here we identify a distinct strain of AAV2 and co-infection with at least one helper virus in blood from US pediatric cases of acute severe hepatitis of unknown etiology. These results are consistent with findings from independent and contemporary studies of acute severe hepatitis in children from Scotland and the UK^12,33^. AAV2 infection may contribute to the pathogenesis and/or severity of the hepatitis, or alternatively, may be a non-pathogenic marker of liver inflammation. Further studies, including serosurveillance and cell culture and animal model studies, are needed to investigate a potential causal role infection from this strain or AAV2 in general may play in this disease.

## METHODS

### Ethics Statement

Remnant clinical samples from cases with acute severe hepatitis were collected and analyzed under “no subject contact” protocols with waiver of informed consent approved by the institutional review boards (IRBs) of University of Alabama, Birmingham, California Department of Public Health, New York State Department of Health, and CDC. Whole blood samples from pediatric controls (age < 18) from Children’s Healthcare of Atlanta were prospectively collected and analyzed under a protocol approved by the Emory IRB (STUDY00000723); parents or guardians of these children provided oral consent for study enrollment and collection and analysis of their samples. Remnant whole blood samples from pediatric controls (age < 18) at University of California, San Francisco (UCSF) were collected, biobanked, and analyzed under a “no subject contact” protocol with waiver of informed consent approved by the UCSF IRB (protocol no. 11-05519). This investigation was reviewed by CDC and conducted consistent with applicable federal law and CDC policy (45 C.F.R. part 46.102(l)(2), 21 C.F.R. part 56; 42 U.S.C. Sect. 241(d); 5 U.S.C. Sect. 552a; 44 U.S.C. Sect. 3501).

### Sample Collection

All acute hepatitis cases in this study were categorized as severe as all patients were hospitalized with acute elevation in liver enzymes, asparate aminotransferase (AST) or alanine aminotransferase (ALT), and one or more of the following symptoms on presentation: nausea, vomiting, jaundice, generalized weakness, and abdominal pain. Acute severe hepatitis cases from California were selected among those sent to the California Department of Public Health for typing and whole-genome sequencing after testing adenovirus positive, depending on sample availability. For cases from Alabama, Florida, Illinois, North Carolina, and South Dakota, samples from PUIs were collected and sent to the CDC for further diagnostic testing. A person under investigation (PUI) was defined as a person <10 years of age with elevated (>500 U/L) aspartate aminotransferase (AST) or alanine aminotransferase (ALT), an unknown etiology for the hepatitis, and onset on or after October 1, 2021^3^. For the Alabama cases (n=8), samples were selected for inclusion into the current study if they had tested positive for adenovirus and if there was sufficient available volume. For the remaining 4 cases from states other than Alabama or California for whom sufficient volume was available, samples were not previously known to be positive for adenovirus. Samples were stored at -80°C until use.

Remnant whole blood samples from controls from UCSF (n=37) were retrospectively biobanked and aliquoted with addition of 2X DNA/RNA Shield (Zymo Research) in a 1:1 ratio by volume and stored at -80°C until use. These controls, all from California, were selected to be geographically similar (located within the same state) to the cases from California. Whole blood samples from controls from Children’s Healthcare of Atlanta (n=8) were obtained from children enrolled into a sample collection protocol following informed consent and assent, as appropriate for age. These controls were all from Georgia and were outpatient volunteers (n=6) or hospitalized patients (n=2), selected to geographically similar (located within a neighboring state with similar demographic characteristics) to the cases from Alabama and Florida^34^. Samples were stored at -80°C until use. Among the controls, 23 (51%) of 45 were collected over the same time frame as the cases (October 1, 2021 to March 28, 2022).

### Nucleic Acid Extraction

Whole blood and plasma samples collected from cases from Alabama were extracted at the Wadsworth Center laboratory using the NucliSENS® easyMAG “specific B” protocol (bioMerieux) according to the manufacturer’s instructions. Samples from cases from Florida, Illinois, North Carolina, and South Dakota were extracted using the Zymo Direct-zol™ DNA/RNA Miniprep Kit (Zymo Research) following the manufacturer’s instructions. Briefly, 200 μL of sample was extracted and total nucleic acid was eluted in 60 μL and stored at -80°C until use.

Samples from cases form California were extracted at the California Department of Public Health. Whole blood samples (200 μL) were extracted using the Qiagen Blood Mini Kit (Qiagen) according to the manufacturer’s instructions. Total nucleic acid was eluted in 100 μL and stored at -80°C until use. Respiratory samples, serum, plasma, and clarified stool suspensions were extracted using the NucliSENS easyMAG instrument (bioMerieux). Briefly, 300 μL of nasopharyngeal swab, serum, or plasma sample or 140 μL of clarified stool suspension was lysed with 1 mL of lysis buffer and incubated for 10 minutes at room temperature, followed by addition of 100 μL magnetic silica and transfer to the instrument to begin the automated extraction. Total nucleic acid was eluted in 110 μL for nasopharyngeal swab, serum, or plasma or 60 μL for stool and stored at -80°C prior to use.

For control samples from California or Georgia extracted at UCSF, total nucleic acid was extracted from the original sample using two different protocols for whole blood and plasma. Whole blood samples (400 μl) that had been pretreated with DNA/RNA Shield (Zymo Research) were extracted using the Quick-RNA Whole Blood Kit (Zymo Research) according to the manufacturer’s instructions. Total nucleic acid was eluted in 15 μl and stored at -80°C until use. Plasma samples (200 μl) were extracted using the Mag-Bind Viral DNA/RNA 96 Kit (Omega Bio-Tek) on a KingFisher Flex instrument (Thermo-Fisher scientific) according to the manufacturer’s instructions. Total nucleic acid was eluted in 100 μl and stored at -80°C until use.

### Viral PCR testing

Samples from California cases were screened for adenovirus using a pan-adenovirus PCR targeting all human adenoviruses^35^ and/or a group F adenovirus (HAdV-40 / HAdV-41) real-time PCR^36^. A cycle threshold cutoff of ≤40 was used to call a positive result by PCR. PCR and Sanger sequencing of the HAdV hexon gene targeting hypervariable regions 1-6^37^ were performed on all adenovirus positive samples with purified PCR products sequenced in-house or sent to an outside laboratory (Sequetech, Mountain View, CA) for sequencing. Sanger sequences were assembled and edited in Sequencher 5.2.4 (Gene Codes) and analyzed using the nucleotide BLAST aligner^38^. Samples from cases outside of California and all control samples were screened for adenovirus using a pan-adenovirus PCR assay^35^ and sequenced using a nested PCR assay targeting HAdV hexon hypervariable regions 1-6, followed by Sanger sequencing^39^. PCR testing for detection of CMV, EBV, and HHV-6 was performed as previously described^40^.

### Viral Enrichment Based Targeted Sequencing

For cases from California and Alabama and all controls, viral enrichment followed by targeted sequencing of viral genomes was performed using custom spiked primers designed to target HAdV-41 or adeno-associated virus (AAV1-AAV8) genomes. Spiked primers were designed using the metagenomic sequencing with spiked primer enrichment (MSSPE) algorithm^17^ as follows. For HAdV-41, 22 representative HAdV-41 genomes were aligned using MAFFT, while for AAV, 11 genomes representing AAV1-AAV8 were aligned using MAFFT. Next, automated primer design was performed by running the MSSPE algorithm with the following parameters: `kmer size=25, segment window=500, e-value=0.1, dS=2, dG=-9000`. The algorithm generated 219 AdV and 150 AAV spiked primers **(Extended Data Table 3)**. Libraries were prepared using NEBNext Ultra II DNA Kit (New England Biolabs). Briefly, adapters were ligated to the purified cDNA (1:25 diluted adaptor), followed by PCR library amplification using the spiked primers and barcoding using NEBNext Multiplex Oligos set 1 (New England Biolabs). Final libraries were quantified using the Qubit Flex instrument (Invitrogen) with the dsDNA HS Assay Kit (Invitrogen). Libraries were pooled and sequenced on a NextSeq 550 Sequencing System using 300 base pair (bp) single-end sequencing. Negative template controls were included in every run to monitor for contamination. No contamination from HAdV-41 or AAV2 reads was detected in the negative template control libraries.

For cases from Florida, Illinois, North Carolina, and South Dakota, probe capture target enrichment of viral genomes was performed on metagenomic libraries using the Twist Comprehensive Viral Research Panel (Twist Biosciences), which covers reference genomes of 3153 viruses and 15,488 different strains^41^. Individual DNA or cDNA/RNA libraries were hybridized to CVRP probes according to the manufacturer’s instructions.

Libraries were then barcoded and sequenced on an Illumina MiSeq (250 bp paired-end sequencing) or an Illumina NextSeq (150 bp paired-end sequencing) (Illumina). For plasma sample 14_NC, nucleic acid carrier, either bacteriophage lambda DNA (New England Biolabs) or HeLa total RNA (Thermo-Fisher), was added during the library preparation step according to the manufacturer’s instructions.

### Viral Metagenomic Sequencing and Analysis

Metagenomic DNA and RNA libraries were prepared using the NEBNext Ultra II DNA Library Prep Kit (New England Biolabs) and NEBNext Ultra II RNA Library Prep Kit (New England Biolabs), respectively, according to the manufacturer’s instructions. Libraries were pooled and sequenced on a NextSeq 550 Sequencing System using 150 bp single-end sequencing. Potential contamination was monitored in each run by processing negative water and controls in parallel with samples.

Sequencing data from all cases and controls were analyzed for viral nucleic acids using SURPI+ (v1.0.7-build.4)^42^, an automated bioinformatics pipeline for pathogen detection and discovery from metagenomic data that has been modified to incorporate enhanced filtering and classification algorithms^20^. A threshold of ≥3 non-overlapping reads was used for calling a positive virus detection^20^.

### Phylogenetic Analysis

Multiple sequence alignments were performed using MAFFT algorithm^22^ as implemented in Geneious (version 10.0.9)^43^. Nucleotide and amino acid phylogenetic trees were inferred using a maximum likelihood method with ultrafast bootstrap approximation as implemented in IQ-TREE (version 1.6.1)^23^ using 1000 bootstrap replicates. Trees were visualized using FigTree (version 1.4.4)^44^.

### Viral Genome Assembly and Analysis

Binary base call (BCL) files generated by Illumina sequencers were simultaneously demultiplexed and converted to FASTQ files using bcl2fastq (version 2.20.0.422). A custom script was used to assemble HAdV and AAV2 genomes as follows. Briefly, raw FASTQ reads were filtered using BBDuk (version 38.87)^45^ for removal of adapters, primer sequences, and low-quality reads, and then HAdV-41 or AAV reads were identified by Bowtie2^46^ alignment (parameters: `-D 20 -R 3 -L11 -N 1`) to a reference database consisting of 1,395 HAdV-41 or 3,600 AAV partial sequences / genomes, respectively. These aligned reads were then mapped to the HAdV-41 reference genome (accession DQ315364.2) or consecutively to AAV genomes 1-8. For all AAV genomes, the assembly with the highest breadth of coverage corresponded to the AAV2 reference genome (accession number NC_001401.2). Consensus assemblies were generated using iVar^47^ (parameters: `-t 0.5-m 1`). AAV consensus genomes were further analyzed for shared mutations in the nucleotide and translated nucleotide (amino acid) sequences relative to the AAV2 reference by performing a multiple sequence alignment using the MAFFT algorithm^22^, followed by visualization of the alignment using Geneious software (version 10.0.9)^43^.

### Statistical Analysis

Statistical analyses were performed using the Python scipy package (version 1.5.2)^48^ and rstatix package (version 0.7.0) in R (version 4.0.3)^49^. Fisher’s Exact Test was used to assess the association between variables. All statistical tests were conducted as two-sided at the 0.05 significance level.

### Data Visualization

Plots were generated using matplotlib (version 3.3.2), seaborn (version 0.11.0) and plotly (version 5.6.0) packages in Python software (version 3.7.12), Jupyter notebook (version 6.1.4), RStudio (version 1.4) and Adobe Illustrator (version 26.4.1) software.

### Data Availability

All data produced in the present study are available upon reasonable request to the authors.

## Data Availability

All data produced in the present study are available upon reasonable request to the authors.

## ACKNOWLEDGEMENTS

We thank the public health officers who provided samples and patient metadata in support of this study: Judy Kauerauf and Lori Saathoff-Huber, Illinois Department of Public Health; Sylvia Bunch, Dalton Dessi, Ashley Gent, Charles Panzera, Samantha Vaccaro and Julieta Vergara Aguirre, Florida Department of Health; Lauren DiBiase, University of North Carolina Hospitals; Lana Deyneka, North Carolina Department of Health and Human Services; Rebecca Pelc, North Carolina Department of Public Health; and the North Carolina State Laboratory of Public Health. We thank Hugo Guevara, Tasha Padilla, Estela Saguar, Chelsea Wright, Blanca Molinar, Lisa Moua, and Alice Chen from the Respiratory and Gastroenteric Diseases Section of the Viral and Rickettsial Disease Laboratory, California Department of Public Health. We thank staff members at the UCSF Clinical Laboratories and the UCSF Clinical Microbiology Laboratories for their help in identifying and retrieving patient whole blood samples for biobanking. We thank the Wadsworth Center Advanced Genomics Technology Core for performing the sequencing reactions, and Simon Ogbamikael for performing the hexon gene-sequencing assays. We thank the Genomics group at NBFAC for their efforts in creating the genomic libraries on short notice from the samples they were provided.

This work was funded in part by US Centers for Disease Control and Prevention contracts 75D30121C12641 and 75D30121C10991 (C.Y.C.), BARDA contract 75A50122C00022, and National Institutes of Health (NIH)/National Institute of Child Health and Human Development (NICHD) grant R61HD105618 (C.Y.C. and C.A.R.). This work was also funded in part under Agreement No. HSHQDC-15-C-00064 awarded to Battelle National Biodefense Institute (BNBI) by the Department of Homeland Security (DHS), Science and Technology Directorate (S&T), for the management and operation of the National Biodefense Analysis and Countermeasures Center (NBACC), a Federally Funded Research and Development Center. This manuscript has been authored in part by Battelle National Biodefense Institute, LLC under contract with DHS. The publisher, by accepting the article for publication, acknowledges that the United States Government retains a non-exclusive, paid up, irrevocable, world-wide license to publish or reproduce the published form of this manuscript, or allow others to do so, for United States Government purposes. The funders had no role in the design and conduct of the study; collection, management, analysis, and interpretation of the data; preparation, review, or approval of the manuscript; and decision to submit the manuscript for publication.

The findings and conclusions in this article are those of the authors and do not necessarily represent the views or opinions of the Alabama Department of Public Health, California Department of Public Health, California Health and Human Services Agency, Florida Department of Health, Illinois Department of Public Health, New York State Department of Health / Wadsworth Center, North Carolina Department of Public Health, or South Dakota Department of Health, nor do they reflect the official policy or position of the FBI or CDC. The views and conclusions contained in this article should not be interpreted as necessarily representing the official policies, either expressed or implied, of DHS or the U.S. government. DHS does not endorse any products or commercial services mentioned in this presentation. In no event shall DHS, BNBI or NBACC have any responsibility or liability for any use, misuse, inability to use, or reliance upon the information contained herein. In addition, no warranty of fitness for a particular purpose, merchantability, accuracy, or adequacy is provided regarding the contents of this document. This is publication #22.28 of the FBI Laboratory Division. Names of commercial manufacturers are provided for identification purposes only, and inclusion does not imply endorsement of the manufacturer, or its products or services by the FBI.

## COMPETING INTERESTS

C.Y.C. is a founder of Delve Bio and on the scientific advisory board for Delve Bio, Mammoth Biosciences, BiomeSense, and Poppy Health. He is also a co-inventor on U.S. patent 11380421, “Pathogen Detection using Next Generation Sequencing, under which algorithms for taxonomic classification, filtering, and pathogen detection are used by SURPI+ software. C.A.R.’s institution has received funds to conduct clinical research unrelated to this manuscript from BioFire Inc, GSK, Janssen, MedImmune, Merck, Moderna, Novavax, PaxVax, Pfizer, Regeneron, Sanofi-Pasteur. She is co-inventor of patented RSV vaccine technology, which has been licensed to Meissa Vaccines, Inc. KSG receives research support unrelated to this manuscript from ThermoFisher and has a royalty generating collaborative agreement with Zeptometrix. The other authors declare no competing interests.

## Notes

### Author Declarations

Remnant clinical samples from cases with acute severe hepatitis were collected and analyzed under no subject contact protocols with waiver of informed consent approved by the institutional review boards (IRBs) of University of Alabama, Birmingham, California Department of Public Health, New York State Department of Health, and CDC. Whole blood samples from pediatric controls (age < 18) from Childrens Healthcare of Atlanta were prospectively collected and analyzed under a protocol approved by the Emory IRB (STUDY00000723); parents or guardians of these children provided oral consent for study enrollment and collection and analysis of their samples. Remnant whole blood samples from pediatric controls (age < 18) at University of California, San Francisco (UCSF) were collected, biobanked, and analyzed under a no subject contact protocol with waiver of informed consent approved by the UCSF IRB (protocol no. 11-05519). This investigation was reviewed by CDC and conducted consistent with applicable federal law and CDC policy (45 C.F.R. part 46.102(l)(2), 21 C.F.R. part 56; 42 U.S.C. Sect. 241(d); 5 U.S.C. Sect. 552a; 44 U.S.C. Sect. 3501).

